# AI-Enhanced Segmentation and Scoring of Histopathological Growth Patterns in Resected Colorectal Liver Metastases

**DOI:** 10.64898/2026.10.21.26366115

**Authors:** Zhen Qian, Cornelis Verhoef, Faber Pas, Michael A. de Leeuw, Johannes H.W. de Wilt, Iris D. Nagtegaal, Michael Doukas, Peter Vermeulen, Stefan Klein, Dirk J. Grünhagen, Martijn P.A. Starmans

**Author notes:** **Corresponding author** Dr. Martijn P. A. Starmans, Assistant Professor & PI Artificial Intelligence for Integrated Diagnostics (AIID) focused on Medical Imaging in Oncology.

## Abstract

Histopathological growth patterns, predominantly the presence of an encapsulated rim, serve as powerful prognostic biomarkers in colorectal liver metastases. However, conventional visual estimation is limited by interobserver variability, human rounding bias, and categorical thresholds. We aimed to develop and externally validate a deep-learning segmentation framework that produces continuous, spatially resolved, and objective encapsulation metrics from routine whole-slide images.

In this multicenter study of chemo-naive resected CRLM patients, a modified nnU-Net architecture was developed on 40 pixel-annotated H&E-stained WSIs to segment liver parenchyma, tumor, and the encapsulation interface. Model performance was optimized across spatial resolutions. An automated continuous AI encapsulation score was computed to determine patient-level metrics. The prognostic value for 5-year overall survival was evaluated using restricted cubic spline Cox regression, a log-rank optimized threshold, and Kaplan–Meier survival analyses across a main cohort (Erasmus MC; *n* = 546, 1,910 WSIs) and an independent external validation cohort (Radboud UMC; *n* = 212, 615 WSIs).

The segmentation network achieved optimal tissue boundary delineation at a resolution of 2.0 µm/pixel on the 40 annotated slides (overall foreground DICE: 0.86 ± 0.09; encapsulation DICE: 0.70 ± 0.11). Spline modeling demonstrated that while visual assessment displayed non-linear hazard plateaus, the continuous AI encapsulation score exhibited a linear reduction in mortality risk across the critical intermediate range (25%–75% encapsulation, *p* = 7.81 × 10^−4^). Log-rank optimization identified a primary global threshold at 47.4% (*p* = 3.73 × 10^−8^), which in confirmed significant OS stratification in external validation (*p* = 0.018). A secondary threshold was found at 69% (*p* = 2.93 × 10^−6^) that captured high-encapsulation favorable-risk patients matching the current manual 100% Rotterdam cut-off.

Our deep-learning model transforms qualitative HGP grading into a continuous, explainable, and fully automated prognostic biometry. By eliminating observer subjectivity and capturing fine-grained risk gradients, the model establishes a scalable, standardized foundation for CRLM risk stratification on digital pathology.

## Introduction

Colorectal liver metastases (CRLMs) display three main canonical histopathological growth-patterns (HGPs): encapsulated, replacement, and pushing, distinguished by their tumour-liver interface architecture. HGPs of CRLMs are proven in multiple retrospective cohort studies to be biomarkers closely related to prognosis.^1^ Over the past decades, different retrospective studies have applied varying definitions and cut-offs to categorize these growth patterns, leading to substantial variability in visually estimated scoring and interpretation.^1–6^

The currently regarded main consensus of 2022, known as the Rotterdam cut-off, differentiates CRLM patients whose metastases are completely (i.e. 100%) encapsulated by a fibrous rim versus non-encapsulated (i.e. <100% encapsulated).^1-7^ The other older consensus of 2017 differentiates between a predominant encapsulated group and predominant non-encapsulated group.^4^ Both methods require visual estimation of the percentage-wise encapsulation score of the tumour rim by expert pathologists.^1^ Recently, Moro et al. (2023) proposed a continuous, proportion-dependent risk gradient, identifying an intermediate-risk subgroup (33%-99% encapsulation) questioning the visual estimation scoring system.^8^ Their method relies on using a labour-intensive and expert-driven manual pixel-wise annotation of the tumour rim, and has not been externally validated. These manual diagnostic techniques are inherently unsuitable for large-scale (validation) studies or integration into routine pathology workflows.

To overcome this challenge, there is a need for a reproducible, labour-efficient, quantitative, and precise alternative. Whole-slide image (WSI) digitisation has enabled the rise of artificial intelligence (AI) tools, which are uniquely suited to handle the complexity and scale of histopathological data. While a previous AI method successfully classified slides as 100% encapsulated versus non-encapsulated (<100% encapsulated), thereby addressing the challenge of reproducibility and scalability, it only used these two classes and thus fell short in capturing the biological nuance of the tumour-stroma interface and limiting explainability, ultimately reducing complex spatial heterogeneity to a single probability score.^9^

We aimed to create an AI-driven segmentation framework that produces continuous, spatially resolved maps of the tumour(−stroma)-liver interface and HGPs on WSIs of colorectal liver metastases, preserving its full morphological complexity. Our proposed method delivers an automated encapsulation metric that substantially improves the speed and consistency demanded by high-throughput clinical pathology workflows. Besides comparison to manual annotations, we internally and externally validate our approach by relating our AI-derived HGP score directly to survival.

## Methods

This investigation was conducted in accordance with the REporting recommendations for TUmour MARKer prognostic studies (REMARK). Ethical approval was secured from both the Erasmus University Medical Centre Medical Ethics Committee (MEC-2018-1743), which waived the need for renewed informed consent, and the Radboud University Medical Centre Ethics Committee (MEC 2015-1637). The study was performed in compliance with the Declaration of Helsinki.

### Datasets

We included two peri-CRLM-operative chemo-naïve cohorts of resected CRLM. The main cohort consisted of 546 consecutive patients, with in total 1910 haematoxylin-and-eosin (H&E) WSIs, resected between 2000-2019 at Erasmus MC (Rotterdam, the Netherlands). The external validation cohort contained 212 patients, with in total 615 WSIs, of Radboud UMC (Nijmegen, the Netherlands) processed during the same calendar period.

Formalin-fixed paraffin-embedded sections (thickness 4–5 µm) were stained with standard H&E and digitised on a 3DHISTECH Pannoramic P1000 scanner equipped with a Plan-Apochromat 20×/0.8 NA objective and an Adimec QUARTZ Q-12A180 CMOS camera (pixel size = 0.24 µm). WSIs were stored in MIRAX™ pyramidal format to enable multi-resolution access.

### HGP assessment and manual segmentation

The HGPs as visual percentages of all WSIs were determined by a team of expert senior pathologists (P.V. and M.D.) according to the international consensus guidelines.^1^

Forty slides of the main cohort (randomly selected, balanced mixture of 100% encapsulated, mixed, or non-encapsulated) of different patients were selected for full manual pixel-wise segmentation (Figure 1A). Pixel-wise ground-truth masks of healthy liver, tumour, encapsulated rim, and background were manually annotated in QuPath (Figure 1B) by an MD PhD researcher (Z.Q.) trained by senior pathologists (P.V. and M.D.).

**Figure 1.**
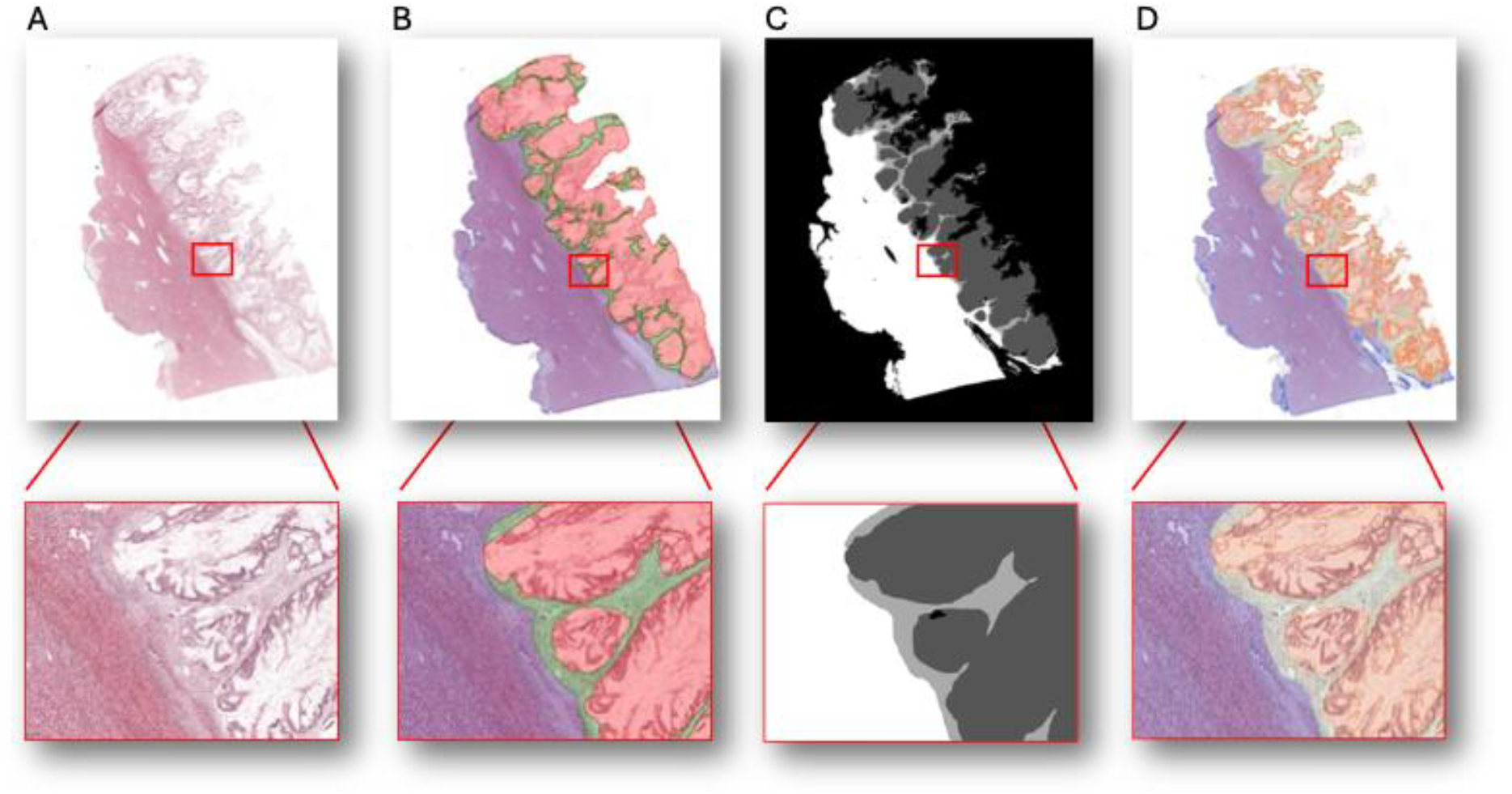
AI-enhanced multi-class tissue segmentation at the colorectal liver metastasis interface. **(A)** Original hematoxylin and eosin (H&E)-stained histological tissue section showing a colorectal liver metastasis and the surrounding liver parenchyma. **(B)** Ground-truth expert manual annotations denoting the segmented tissue compartments: liver parenchyma (purple/blue), tumor tissue (red), and the fibrous tissue/encapsulation rim (green). **(C)** AI model segmentation mask showing class delineations (white: liver tissue; dark grey: tumor; light grey: encapsulation rim; black: background). **(D)** Qualitative AI segmentation overlay on the original H&E image showing fine-grained delineation of the tumor-liver interface.

### Model construction and output generation

We adopted the self-configuring deep learning nnU-Net framework as the basis for our biomedical segmentation model.^10^ As the framework was originally designed for radiological images, several improvements were made.

As preprocessing, artifact removal using H&E Otsu colour thresholding, and Reinhard colour stain normalisation were implemented to improve the quality and interpretability of the WSIs for the neural network.^11–13^ To handle the large size of the WSIs, we adopted a commonly used patch-based strategy, extracting 512 × 512-pixel tiles. To address disproportionate representation of tissue types during training, class-balanced tile harvesting was done to equalise numbers of tumour, capsule, and liver patches using the Whole-Slide-Data package.^14^ Background patches were excluded due to their low relevance. The region selection probability was set proportional to the area of a region to ensure no disproportionate prevalence of small regions. Binary morphological opening and closing removed spurious islands (< 32 × 32 px).

As postprocessing, we introduced two tissue-aware rules to discard trivial HGP irrelevant label combinations: 1) removal of isolated encapsulated areas within the tumour; and 2) removal of isolated islands outside the main slide region.

Segmentation quality was evaluated using five-fold stratified cross-validation on the 40 annotated whole slide images using the DICE coefficient. To evaluate the trade-off between high resolution with more detailed cellular information and a lower resolution with more contextual information, experiments were performed at resolutions of 0.5, 1.0, 2.0, 4.0, 8.0, and 16.0 µm/pixel. Afterwards, a model at the best performing resolution was trained on all 40 development patients and applied to the remaining WSIs. For these 40 patients, the test segmentation results from the cross-validation were used for further analyses.

Models were trained on GPU compute nodes of the Snellius supercomputer (SURF), each equipped with NVIDIA A100 GPUs (40GB variant). During training, a batch size of 6 was used due to memory constraints. Per epoch, 200 batches were used for training and 50 for validation, for a total of 250 epochs.

### Encapsulation score calculation

To calculate the encapsulation percentage from the resulting segmentations, we formulated the slide continuous encapsulation score (*S*), which was designed to most accurately replicate the pathologists’ procedure. The slide continuous encapsulation score was defined by the following equation:

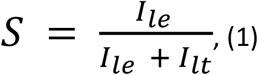

where *I*_*le*_ is the cumulative pixel length of the liver–encapsulation interface and *I*_*lt*_ the cumulative pixel length of the liver-tumor interface of one WSI.

Afterwards, to obtain the patient-level percentage score, the final encapsulation score (*S*_final_) was calculated by taking the weighted average of the slide-level scores (*S*_i_) based on their relative border pixel counts, as follows:

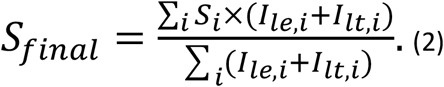

And after simplification:

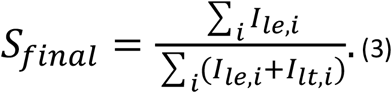

This ensured that slides with longer tumour-liver interfaces (e.g. larger metastases) contribute proportionally more to the overall patient encapsulation score. With the manual visual scoring method this correction was not possible as each slide of each metastasis would have the same weight to *S*_final_.

### Statistical analysis

Survival analysis was first performed on the main cohort patients to evaluate the prognostic value of different HGP encapsulation scoring. Overall survival (OS), defined as the time from CRLM surgery to death from any cause, was censored at 60 months.

Survival distributions were estimated using the Kaplan-Meier method, and differences between strata were assessed via the log-rank test. To compare to cut-offs of the different previously proposed manual HGP encapsulation scores, the optimal cut-off value for the AI-derived slide continuous encapsulation score (*S)* was determined utilizing a grid search for the cut-off yielding the lowest log-rank p-value and using a bootstrap analysis with 10 000 iterations to compute 95% confidence interval (CI). This score was then externally validated on the external validation cohort.

The functional relationship between encapsulation percentage, both for the AI-derived and manual score, and mortality risk was modeled using Cox proportional hazards regression. To account for non-linearities, we employed restricted cubic splines with three degrees of freedom. Model fit and deviations from linearity were assessed using the Analysis of Variance (ANOVA). Inter-method agreement between pathologist manual scoring and AI-model classification was visualized using Sankey diagrams. Statistical significance was defined as a two-sided *p < 0*.*05*. All statistical analyses were conducted in R version 4.5.1 (https://www.r-project.org/).

## Results

### Baseline characteristics

Characteristics of both cohorts are shown in Table 1. The median number of resected CRLMs was one [IQR, 1-2] in both cohorts, with a median largest lesion size of 2.9 cm [2.0-4.0] and 2.6 cm [2.0-4.0], respectively. The disease-free interval (DFI) between primary tumour and liver metastasis was longer in the main cohort (11.0 months [0.0-22.0]) than in the validation cohort (6.5 months [0.0-19.0]). Preoperative CEA levels were comparable across both cohorts (median 11.0 μg/L [4.2-29.0] vs 10.0 μg/L [4.0 – 29.0]).

**Table 1:** Baseline characteristics of the main and validation cohort.

|  | Missing (%) | Value | Main cohort | Validation cohort |
| --- | --- | --- | --- | --- |
| All patients |  |  | 546 | 212 |
| Age at resection of CRLM (median [IQR]) |  |  | 66.00 [59.00, 73.00] | 66.00 [59.00, 72.50] |
| Sex |  | Female | 192 (35.2%) | 74 (35%) |
|  |  | Male | 354 (64.8%) | 138 (65%) |
| ASA Class | 9 (1.6) EMC | I-II | 476 (87.2%) | 176 (84%) |
|  | 2 (0.9) Radboud | III-IV | 61 (11.2%) | 34 (16%) |
| Primary tumour location | 15 (2.7) EMC | Right sided | 107 (19.6%) | 53 (25%) |
|  | 4 (1.9) Radboud | Left sided or rectal | 424 (77.6%) | 155 (75%) |
| T-stage | 10 (1.8) EMC | pT 0-2 | 100 (18.3%) | 37 (18%) |
|  | 7 (3.3) Radboud | pT 3-4 | 436 (79.9%) | 168 (82%) |
| N-stage | 12 (2.2) EMC | N0 | 224 (41.0%) | 77 (36%) |
|  | 1 (0.5) Radboud | N+ | 310 (56.8%) | 134 (64%) |
| Number of treated CRLMs (median [IQR]) |  |  | 1.00 [1.00, 2.00] | 1.00 [1.00, 2.00] |
| Largest CRLM in cm (median [IQR]) |  |  | 2.90 [2.00, 4.00] | 2.60 [2.00, 4.00] |
| DFI in months (median [IQR]) |  |  | 11.00 [0.00, 22.00] | 6.50 [0.00, 19.00] |
| Preoperative CEA in µg/L (median [IQR]) |  |  | 11.00 [4.20, 29.00] | 10.00 [4.00, 29.00] |
| Resection margin involved | 5 (0.9) EMC | No | 483 (88.5%) | 194 (91.5%) |
|  |  | Yes | 58 (10.6%) | 18 (8.5%) |
| Extrahepatic disease |  | No | 494 (90.5%) | 206 (97.2%) |
|  |  | Yes | 52 (9.5%) | 6 (2.8%) |

Resection margins were microscopically involved in 10.6% of the main cohort and 8.5% of the validation cohort. Extrahepatic disease was present in 9.5% and 2.8% of patients respectively. Missing data was overall minimal across all variables (<3% for all fields).

### Model optimisation and calculations

A five-fold cross validation on 40 annotated slides revealed that segmentation performance improved with decreasing image resolution until stabilizing within the 2–4 µm/pixel range (Table 2). At the highest magnification (0.5 µm/pixel), overall model performance was modest (total average DICE overall all classes = 0.77 ± 0.09), with the encapsulation class showing the greatest variability (0.66 ± 0.13). Performance increased consistently at lower magnifications, reaching a plateau at 2–4 µm/pixel, where the total DICE stabilized around 0.86–0.87 ± 0.09. At 8 µm/pixel and coarser resolutions, encapsulation performance deteriorated markedly (0.55 ± 0.27 at 8 µm/pixel; 0.47 ± 0.25 at 16 µm/pixel), whereas tumor and liver segmentation performance remained high (0.94 and 0.98 mean DICE).

**Table 2.** Segmentation performance across varying image resolutions.

| Resolution<br>($\mu\text{m}/\text{pixel}$) | Encapsulation | | | |
| --- | --- | --- | --- | --- |
|  |  | Tumour | Liver | Total Foreground DICE |
| 0.5 | $0.66 \pm 0.13$ | | | |
| 1.0 | $0.70 \pm 0.14$ | $0.81 \pm 0.08$ | $0.85 \pm 0.02$ | $0.77 \pm 0.09$ |
| 2.0 | $0.70 \pm 0.11$ | $0.89 \pm 0.10$ | $0.94 \pm 0.02$ | $0.84 \pm 0.10$ |
| 4.0 | $0.70 \pm 0.12$ | $0.92 \pm 0.10$ | $0.97 \pm 0.02$ | $0.86 \pm 0.09$ |
| 8.0 | $0.55 \pm 0.27$ | $0.93 \pm 0.10$ | $0.98 \pm 0.02$ | $0.87 \pm 0.09$ |
| 16.0 | $0.47 \pm 0.25$ | $0.94 \pm 0.10$ | $0.98 \pm 0.03$ | $0.82 \pm 0.17$ |

Dice similarity coefficients (presented as mean ± 95% confidence interval (CI)) for semantic segmentation of encapsulation, tumour, and liver tissue compartments across whole-slide image resolutions ranging from 0.5 to 16.0 $\mu\text{m}/\text{pixel}$ , including the mean total foreground Dice score of all three tissue types.

Based on this analysis, a resolution of 2 µm/pixel was selected for all subsequent experiments, offering a good trade-off between segmentation accuracy, morphological fidelity, and computational efficiency. Figure 1C-D shows an example result of the segmentation model output.

### Survival Analysis and Cut-off Validation

Spline-based Cox proportional hazards regression analysis utilizing 4-knot restricted cubic splines (3 degrees of freedom; reference baseline HR=1.0 at 0% encapsulation; *N = 518, events = 274*; Figure 2) confirmed a non-linear relationship between encapsulation percentage and survival. Both the visual and AI-derived encapsulation score displayed an initial hazard ratio plateau at low percentages, followed by a decline as encapsulation increased, and reaching the lowest hazard ratio at the highest percentage of encapsulation. In the middle range of 25% to 75% encapsulation, the AI model (Figure 2B) showed a nearly perfect linear relationship with mortality risk (*p=0*.*000781*). Contrarily, visual pathology estimation (Figure 2A) displayed marked non-linearity (non-linearity test *p=0*.*00665*), characterized by an initial risk plateau with minimal protective benefit across lower values (0%-40%) followed by an abrupt, step-like decline beyond the 50% threshold.

**Figure 2.**
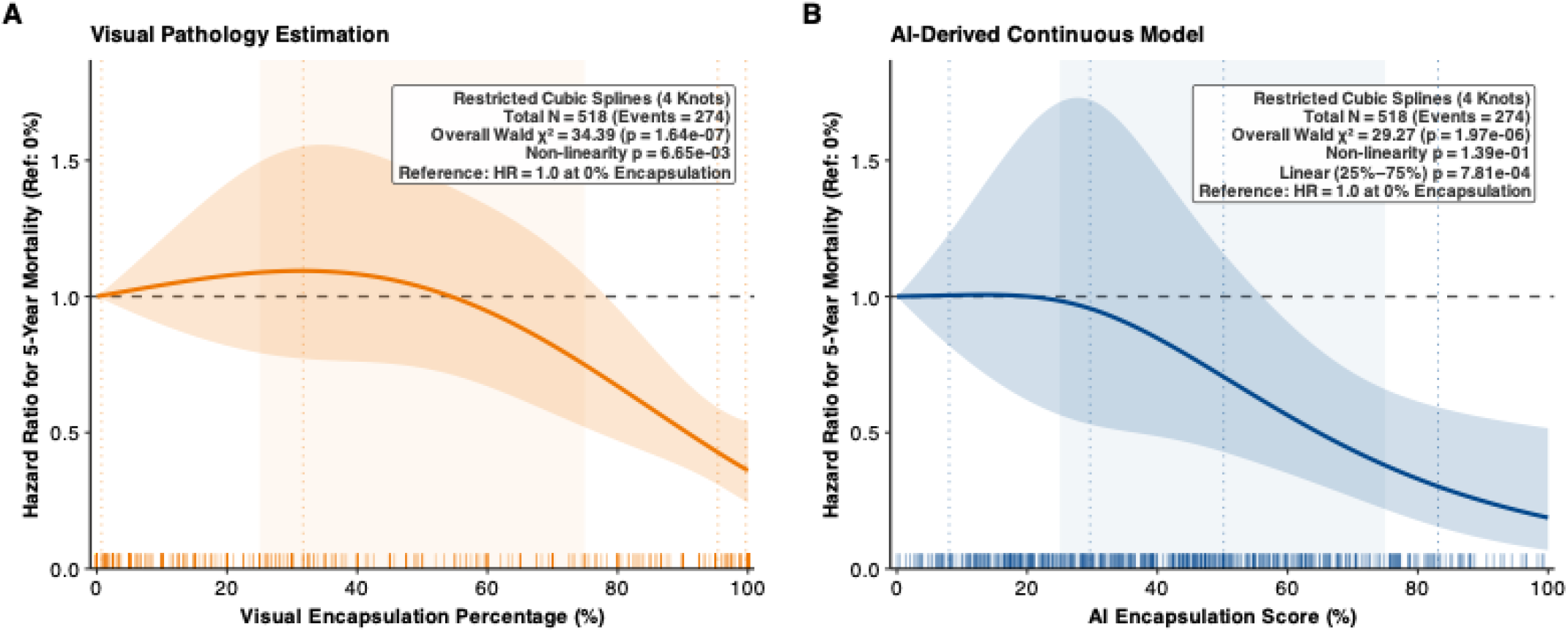
Restricted cubic spline Cox regression of 5-year mortality risk across visual vs. AI continuous encapsulation scores (*N* = 518, events = 274). **(A) Visual pathology scores**. Continuous hazard ratio (HR; solid orange line, 95% CI shaded) modeled with 4-knot splines (reference: HR = 1.0 at 0% encapsulation, horizontal dashed line; knot locations dotted). Visual scoring exhibited marked non-linearity with a hazard plateau at low percentages (0%-40%) followed by a steep drop toward complete encapsulation. Rug plots along the *x*-axis denote individual patient distributions. **(B) AI continuous scores**. Continuous HR (solid blue line, 95% CI shaded) scaled to 0–100%. Within the intermediate transition range (25%–75%, vertical shaded band), the AI metric demonstrated a steady, dose-dependent linear hazard reduction. Rug plots along the *x*-axis denote individual patient distributions.

Using the established visually scored Rotterdam cut-off (Figure 3C), patients with a fully encapsulated growth pattern exhibited a profound survival advantage compared to those with non-fully encapsulated patterns (*p=3*.*72×10*^*-9*^). Similarly, the cutoff used before the second edition of the international scoring guidelines of 2022 (>50% visual encapsulation; Figure 3A) also provided significant risk stratification (*p=1*.*04×10*^*-8*^), though with a lower level of statistical significance.

**Figure 3.**
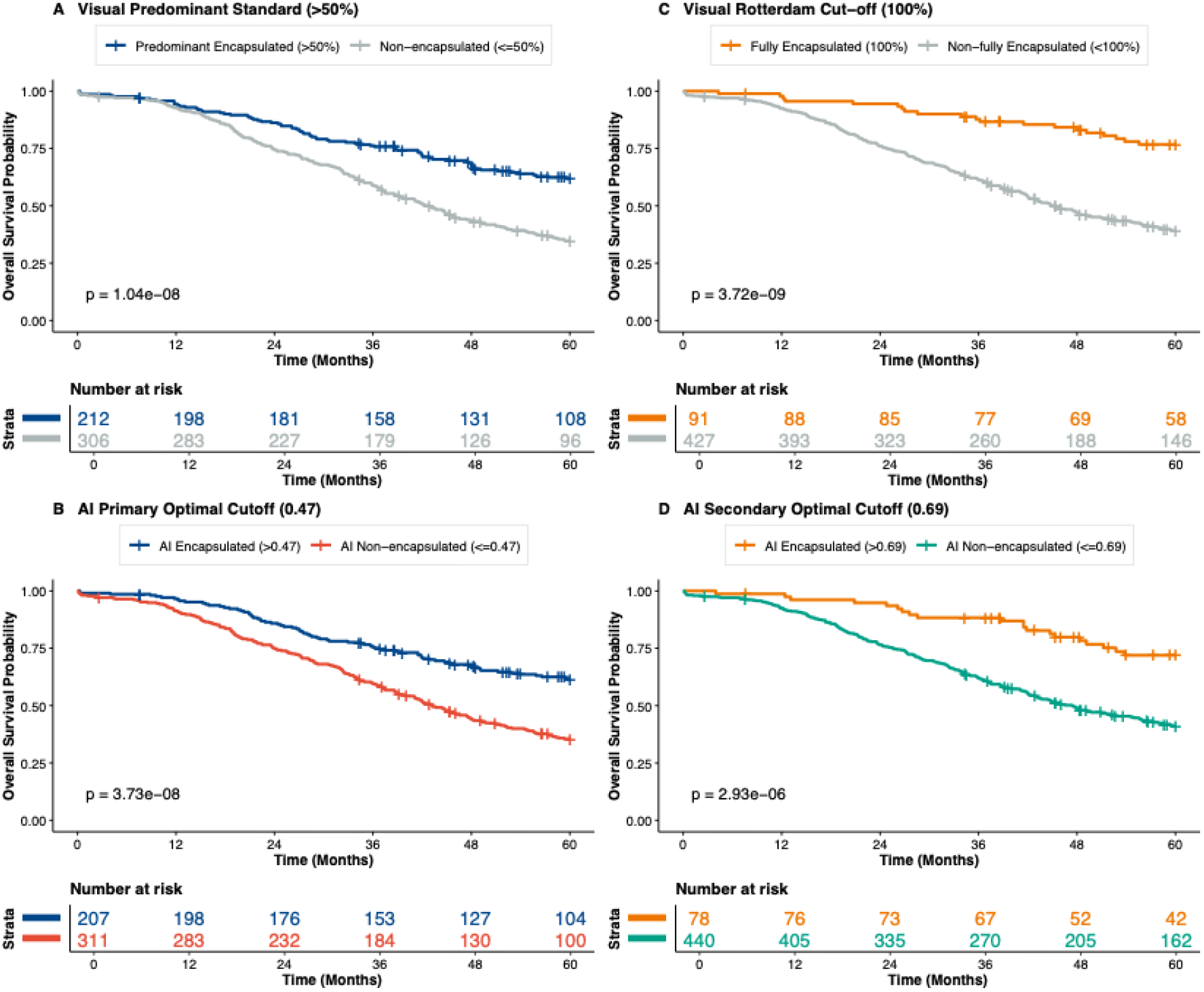
Kaplan–Meier 5-year overall survival analyses comparing visual consensus standards with AI-derived optimal cutoffs (*N* = 518). **(A) Visual Predominant Standard (>50%):** Stratification by the predominant visual cutoff (> 50% vs. ≤50% encapsulation; log-rank *p* = 1.04 × 10^−8^). **(B) AI Primary Optimal Cutoff (0.47):** Stratification by the global optimal AI continuous score cutoff (> 0.47 vs. ≤0.47; log-rank *p* = 3.73 × 10^−8^). **(C) Visual Rotterdam Cutoff (100%):** Stratification according to the current consensus Rotterdam visual cutoff (100% fully encapsulated vs. < 100% non-fully encapsulated; log-rank *p* = 3.72 × 10^−9^). **(D) AI Secondary Optimal Cutoff (0.69):** Stratification by the secondary optimal AI continuous score cutoff (> 0.69 vs. ≤0.69; log-rank *p* = 2.93 × 10^−6^).

Next our automated AI segmentation model was utilized to calculate slide continuous encapsulation score per slide and to determine the value of *S*_final_ for each patient. Using a log-rank optimization approach to identify the threshold that best stratifies patient survival (Figure 4), the optimal cut-off was determined to be 47.4% (*p=1*.*98×10*^*-8*^, Figure 4). This AI-derived cutoff shows high agreement with the predominant cut-off (Figure 5A) and achieved comparable prognostic discrimination in Kaplan-Meier survival analysis.

**Figure 4.**
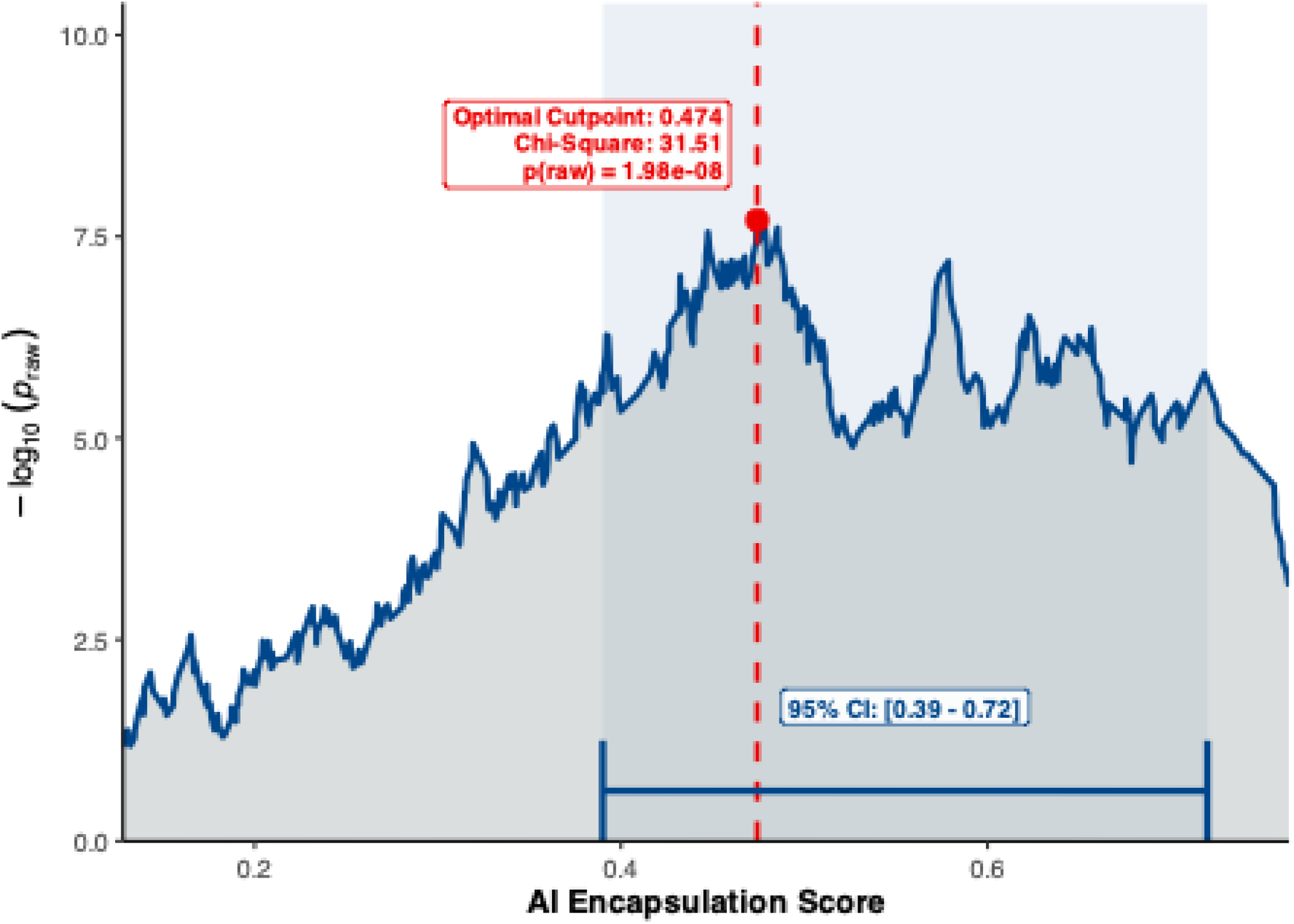
Systematic Log-Rank Grid Search Across the Main Cohort. The vertical axis represents the statistical separation achieved at each AI encapsulation score cutoff as − log_10_(*p*_raw_). The red point and dashed reference line denote the optimal cut-off value corresponding to the maximally selected rank statistic, accompanied by its log-rank χ^2^ and raw *p*-value. The lower bracket in blue indicates the 95% confidence interval estimated using 10.000x bootstrapping. To the right of this area, some additional prominent local maxima can be found, illustrating secondary optimal cutoffs.

**Figure 5.**
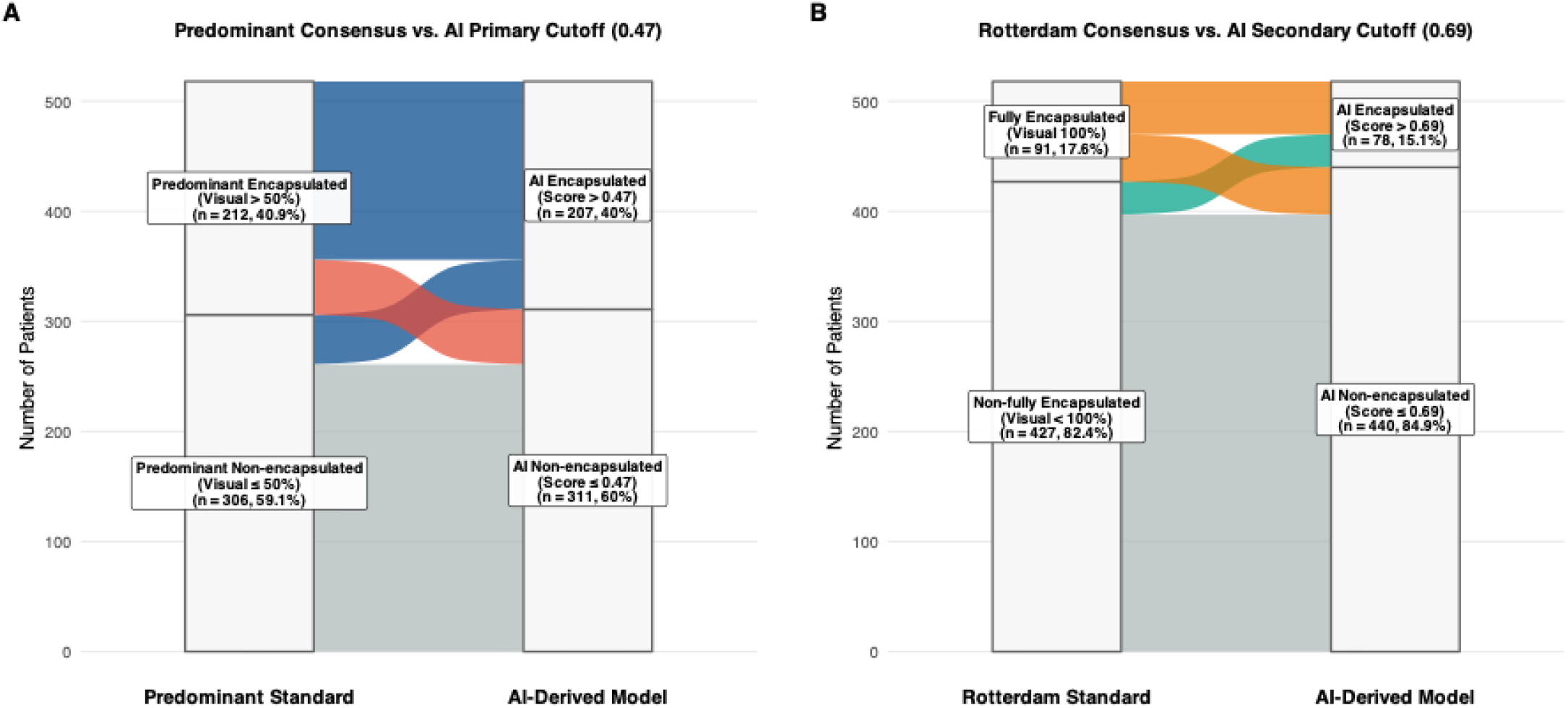
Sankey charts. Sankey charts illustrating the migration and reclassification of patients (*N* = 518) from conventional visual histopathological grading standards (left columns) to objective, AI continuous score thresholds (right columns). Strata blocks denote the total patient count (*n*) and relative proportion within each diagnostic classification. **(A) Predominant consensus standard vs. AI continuous score primary cut-off (0.47)**. **(B) Strict Rotterdam consensus standard vs. AI continuous score secondary cut-off (0.69)**.

The log-rank optimization (Figure 4) showed several other cut-offs with high survival stratification. The value of 0.69 was found to lead to similar groups as the Rotterdam cut-off (Figure 5B), with Kaplan-Meier survival analysis showed similar discriminatory power (Figure 3C-D). This indicates that both previous consensus cut-offs are captured by our AI-derived encapsulation score.

To establish generalizability without the statistical power loss of dichotomization, the prognostic value of the AI score was externally evaluated as a continuous metric in the external validation cohort (*N = 212*; Figure 6). Each 10% increase in AI continuous score was significantly associated with a 12% reduction in 5-year mortality risk (HR = 0.88, 95% CI: 0.78 – 1.00, *p=0*.*044*), closely matching the development cohort (HR = 0.86, 95% CI: 0.82 – 0.91, *p=5*.*08×10-8*). Discriminative performance was preserved externally (Harrell’s C-index = 0.578, 95% CI: 0.508-0.648 vs 0.592, 95% CI: 0.558-0.626 in the main cohort) and outperforming visual scoring (Harrell’s C-index = 0.572, 95% CI: 0.551-0.594). Comparative restricted cubic splines confirmed congruent monotonic hazard trajectories across both centers, with no significant deviation from linearity (*p=0*.*139* in the main cohort, *p=0*.*653* in the external cohort). In cut-off analyses, the optimal cutoff retained significant risk separation in the external cohort (*p=0*.*018;* Figure 7A and 7B), mirroring the visual predominant standard. Conversely, the secondary optimal cutoff restricted the favorable-prognosis group in the external cohort (Figure 7D), but lost statistical significance, highlighting the vulnerability of rigid cutoffs to inter-cohort prevalence differences.

**Figure 6.**
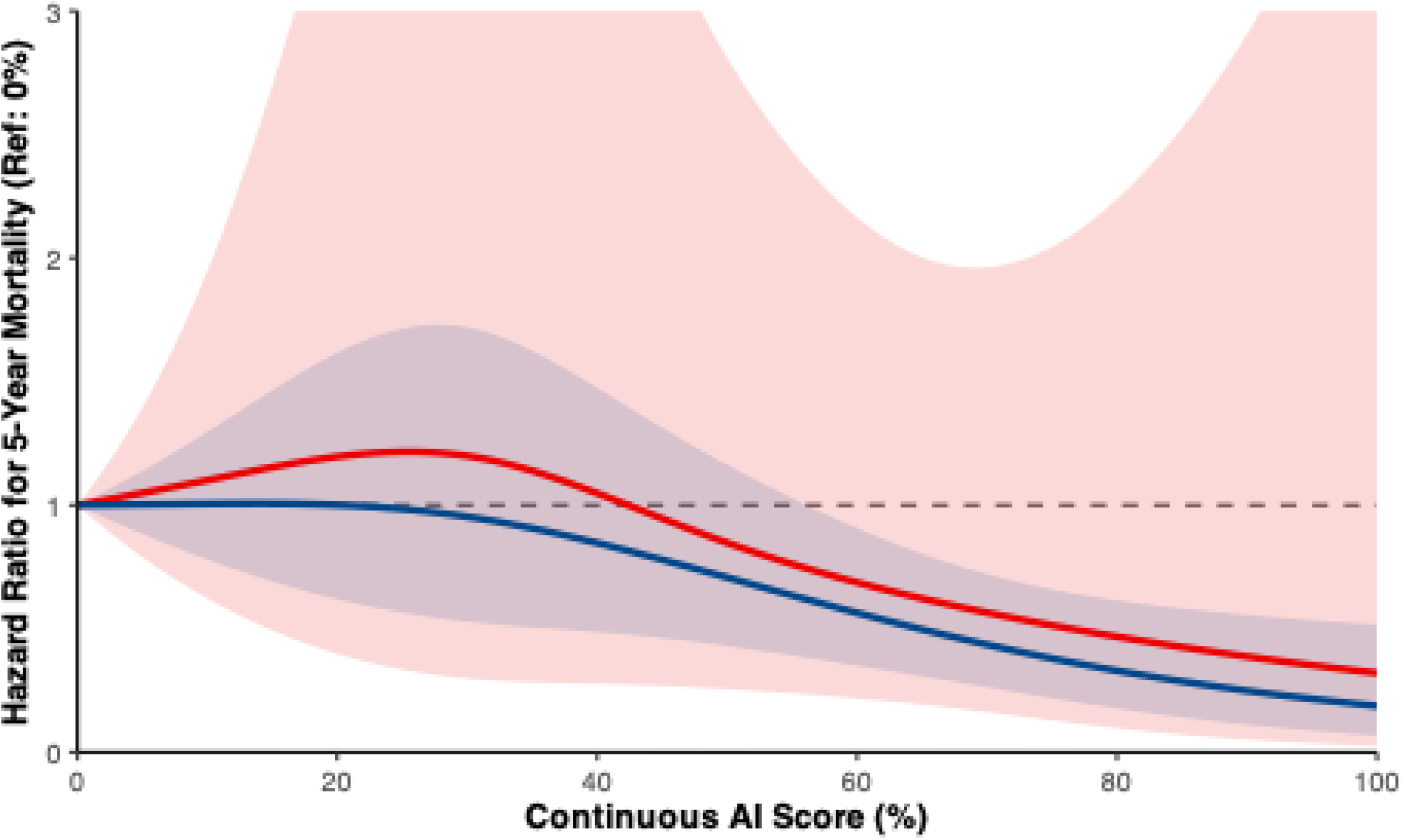
Restricted cubic spline regression of 5-year mortality HR evaluation. Hazard ratios (solid lines) and 95% confidence intervals are shown for the main cohort (blue line) and the external cohort (red line).

**Figure 7.**
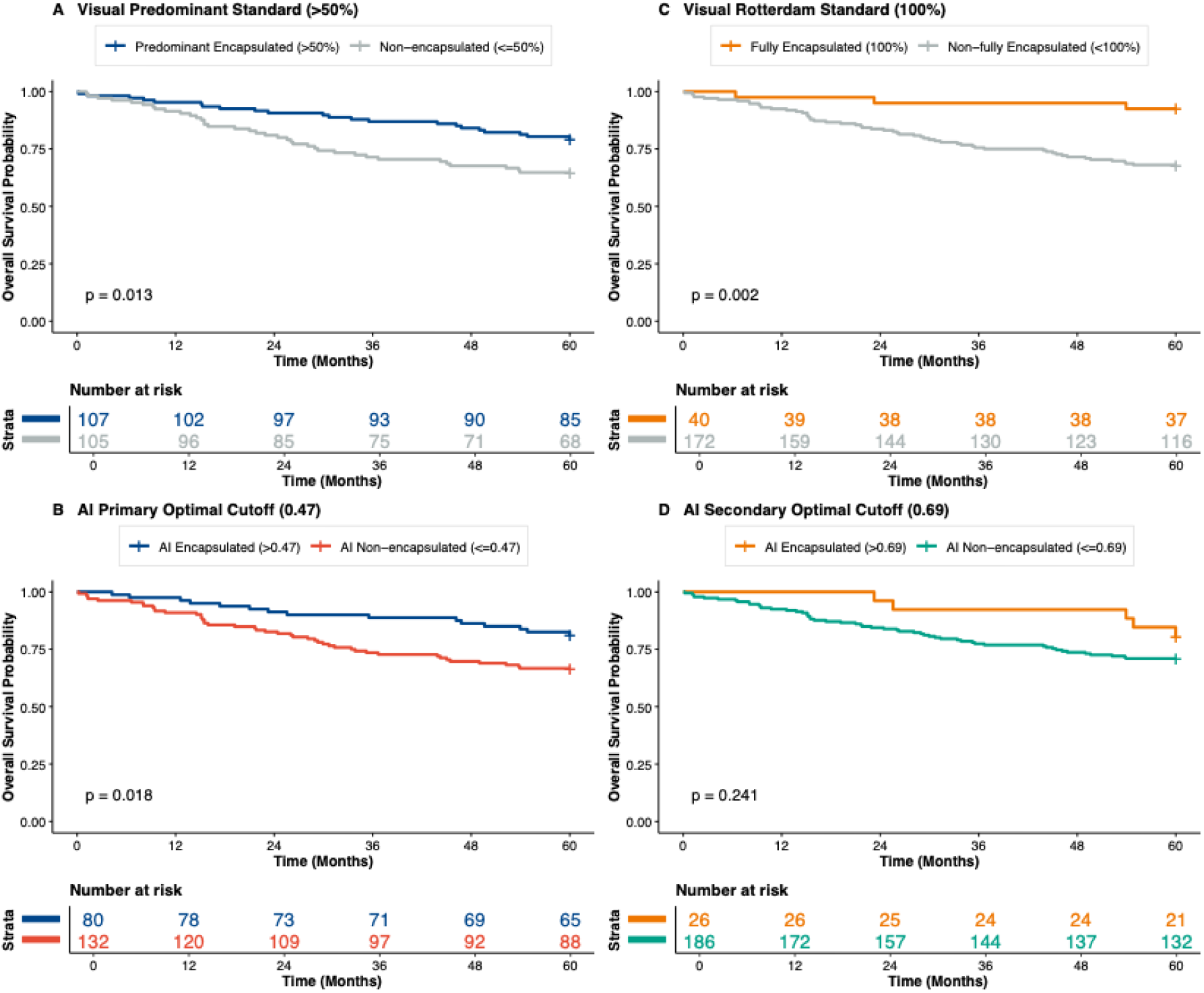
External cohort validation of 5-year overall survival comparing visual consensus standards with AI-derived optimal cutoffs (*N* = 212). **(A) Visual Predominant Standard (>50%):** Stratification by visual estimation using the historical consensus threshold (> 50% vs. ≤50% encapsulation; log-rank *p* = 0.013). **(B) AI Primary Optimal Cutoff (0.47):** Stratification by the continuous AI model evaluated at the primary global threshold (> 0.47 vs. ≤0.47; log-rank *p* = 0.018). **(C) Visual Rotterdam Standard (100%):** Stratification according to the strict visual criteria (100% fully encapsulated vs. < 100% non-fully encapsulated; log-rank *p* = 0.002). **(D) AI Secondary Optimal Cutoff (0.69):** Stratification by the secondary local AI threshold (> 0.69 vs. ≤0.69; log-rank *p* = 0.241). Tick marks indicate censored observations; tables below each panel denote the number of patients at risk through 60 months of follow-up.

## Discussion

This study presents a fully automated AI model that quantifies the histopathological growth patterns of CRLMs through continuous, spatially resolved, and explainable encapsulation segmentation maps. Our model overcomes the limitations of the current visual estimation by pathologists by providing an observer independent, accurate, low-labor, and time-efficient scoring method. Moreover, our results show that our AI-derived encapsulation score provides a more continuous relation with mortality risk and survival than the visual score, while being able to replicate the stratification from the previous visual score consensus cut-offs.

Our findings confirm that encapsulation percentage is a strong prognostic marker for overall survival, irrespective of whether it is visually estimated or AI-calculated. Within both methods and in both cohorts, higher degrees of encapsulation consistently correlated with improved survival outcomes. The proposed AI pipeline offers several methodological advantages that address the inherent limitations of traditional visual HGP scoring. By automatically producing pixel-wise segmentation maps on readily available H&E slides, the model provides spatial explainability that highlights the local tumor-liver interactions underlying global survival outcomes. This transition to full automation removes the requirement for expert further manual input, allow weighted tumor size encapsulation scoring, enabling large-scale application, unbiased standardization, and quality control across cohorts and institutions.

Next to the observed concordance with known visual scoring systems, our AI-derived encapsulation score shows a more granular risk stratification between growth patterns and outcome. Moreover, the AI-calculated encapsulation percentage demonstrates a superior relationship with OS when evaluated via hazard ratios. While manual visual estimation scores are frequently characterized by “plateaus” where survival hazard ratios converge due to potentially different scoring methods over the years, human rounding errors, estimations, the automated model provides a more robust quantitative gradient. Specifically, when analyzing the middle range, the AI-calculated score showed a strong linear correlation with the hazard for survival. This further suggests that the AI-model can capture the biological relationship at the tumor-liver interface with more precision than the manual visual scoring method.

Our study has several limitations. Firstly, the ground-truth masks were generated by a single annotator on slides from one scanner, leaving inter-observer and inter-scanner variability of the segmentation performance untested. However, the survival analysis was conducted on a multi-center dataset with multiple annotators for the HGP score, supporting generalization of our conclusions. Second, patients treated with chemotherapy were excluded, hence no claims can be made regarding the performance of our model in this population. The morphology of the resected specimen could be visually different from our cohort as therapy-induced necrosis, fibrosis and inflammation is inherently underrepresented in chemo-naïve cohorts and could alter the segmentation accuracy of the tumor-liver interface. The exclusion was intentional, as the survival benefit of encapsulation in chemo-exposed metastases is less clearly defined and even debated, and thus there is currently no method to distinguish between non-therapy induced encapsulation/replacement and therapy-induced necrosis and fibrosis/chemoresistance.^4^ Incorporating chemo-exposed slides into model training and explicitly modelling other treatment-related histologic changes, such as acellular mucin pools, necrotic cavities, and reparative fibrosis, could improve morphological generalization.

Our method paves the way for future research on exploring additional HGP-related biomarkers for prognostication besides the encapsulation score. We have on purpose constrained our current analysis to the encapsulation score to closely mimic existing HGP scoring guidelines. However, the current scoring iteration weighs every boundary pixel equally thus potentially penalizing minor irregularity and does not look at the cell decomposition of the border nor the tumor. Morphological features like rim continuity, stromal thickness, and lymphocytes of the encapsulation border and the surrounding area could provide additional survival stratification. By better representing these potentially biologically meaningful spatial characteristics, our model has the potential for even more granular stratification of CRLM patients.

## Conclusion

Our fully automated deep-learning framework for HGP scoring in CRLMs provides an automatic and continuous prognostic metric compared to traditional visual assessment. This model can be seamlessly integrated in the pathology workflow or be an aid in learning HGP scoring, which will become an important factor during the pathology workup of a CRLM as HGP scoring has become an important parameter in the updated International Collaboration on Cancer Reporting datasets.^15^ By eliminating interobserver variability and capturing linear hazard relationships that manual scoring often overlooks, particularly in intermediate encapsulation ranges, our model offers a high-fidelity representation of the tumor-liver interface. The transition from dichotomous, expert-dependent grading to spatially resolved, pixel-wise biometrics establishes a reproducible and scalable standard for HGP implementation. Ultimately, this AI-driven approach facilitates objective risk stratification and provides a foundation for integrating digital pathology into the clinical management of patients with colorectal liver metastases.

## Acknowledgments

This work used the Dutch national e-infrastructure with the support of the SURF Cooperative using grant no. EINF-12583. Martijn Starmans acknowledges funding from the NGF AiNed Fellowship Grants (project number NGF.1607.22.025), which is (partly) financed by the Netherlands Organisation for Scientific Research (NWO).

## Data availability

The script and models used for colorectal liver metastases segmentation in this study are openly available on GitLab at https://gitlab.com/radiology/radiomics/pathology/crlm_segmention under the Apache 2.0 license (commit ID: 583279bf6f69276642c69c2ef5694c8e1ed4fd78).

## Author contributions

**Z.Q**.: Study conceptualization, data annotation, experimental design, data analysis, and original draft preparation.

**C.V**.: Resource provision, study conceptualization, co-supervision and manuscript editing.

**F.P**.: Technical pipeline development, data analysis, and manuscript editing.

**M.A.d.L**.: Technical pipeline development, data analysis, and manuscript editing.

**J.H.W.d.W**.: Resource provision and manuscript editing.

**I.D.N**.: Resource provision and manuscript editing.

**M.D**.: Pathological review and manuscript editing.

**P.V**.: Pathological review and manuscript editing.

**S.K**.: Study conceptualization, co-supervision, and manuscript drafting.

**D.J.G**.: Study conceptualization, co-supervision, and manuscript drafting.

**M.P.A.S**.: Study conceptualization, co-supervision, and manuscript drafting.

## Tables

Dice similarity coefficients (presented as mean ± 95% confidence interval (CI)) for semantic segmentation of encapsulation, tumour, and liver tissue compartments across whole-slide image resolutions ranging from 0.5 to 16.0 µm/pixel, including the mean total foreground Dice score of all three tissue types.

